# Acetate, a fibre-derived gut metabolite, is associated with reduced cardiovascular disease risk in females with early menopause

**DOI:** 10.64898/2026.02.10.26346040

**Authors:** Chaoran Yang, Shinichi Namba, Koichi Matsuda, Yukinori Okada, BioBank Japan Project, Nele Taba, Kreete Lüll, Lisa Moran, Amanda Vincent, Francine Z. Marques

## Abstract

**Background and Aims:** Estrogen deficiency and testosterone excess substantially increase cardiovascular disease risk in females. Dietary fibre and its microbial by-products, short-chain fatty acids, have cardioprotective effects. We aim to investigate whether plasma acetate – the most abundant short-chain fatty acid – is associated with improved cardiovascular outcomes in females with altered sex hormone profiles.

**Methods:** This cohort study included 105,563 female participants from the UK Biobank and Biobank Japan with up to 10 years of follow-up. The primary outcome was major adverse cardiovascular event (MACE) in relation to early menopause (surrogate for estrogen insufficiency) and plasma free testosterone. Proteomics profiling explored underlying molecular pathways.

**Results:** Higher plasma acetate was associated with lower 10-year MACE incidence (HR=0.887, *p*=0.002) in the UK Biobank. Acetate levels above the median attenuated the high MACE risk associated with early menopause (HR=1.155, *p*=0.075) compared with lower acetate levels (HR=1.431, p<0.001), further supported by a negative early menopause × acetate interaction on MACE incidence (HR=0.894, *p*=0.037). This mitigation pattern was replicated in Biobank Japan (above median: HR=1.350, *p*=0.067; below median: HR=1.423, *p*=0.028). The top acetate quartile (>27.1g fibre/day) attenuated the early menopause-associated MACE risk. Proteomics implicated the underrepresentation of pro-inflammatory pathways. High acetate also attenuated MACE risk associated with elevated free testosterone, although the interaction term was not significant.

**Conclusions:** Higher plasma acetate mitigated cardiovascular risk in females, particularly in those with early menopause, potentially by modulating pro-inflammatory pathways. These findings support recommendations for higher dietary fibre intake as a CVD prevention strategy for females with early menopause.

**Translational Perspective:** Plasma acetate is the key product of gut microbial fermentation of fibre. Using two cohort studies, totalling 105,563 female participants followed for 10-years from two independent biobanks with different ethnicities, higher plasma acetate was significantly associated with lower cardiovascular disease risk, especially for those with early menopause. Higher microbial acetate protects against cardiovascular disease in females, with greater benefits in those with hormone related cardiovascular vulnerability. These findings may inform the design of clinical trials and guidelines, supporting a personalised nutrition strategy based on hormonal profile.

## Introduction

Cardiovascular disease (CVD), including ischaemic heart disease and stroke, is the leading cause of death worldwide^1^ and is expected to remain so until 2050.^2^ However, females have disproportionately worse CVD outcomes than males due to inequalities in prevention, diagnosis, and treatment.^3,4^ Although premenopausal females exhibit a lower incidence of CVD compared to males, there is a marked increase in CVD in females after menopause,^5–7^ with dysregulation of sex hormone levels being an essential contributor.^6^ Notably, altered sex hormone production is associated with significantly increased CVD risk in women.^8,9^ For instance, early or premature menopause (menopause before age 45 or 40 years, respectively^10^), which is characterised by oestrogen deficiency, is associated with a 24-87% higher CVD hazard ratio (HR).^9,11^ Moreover, Polyendocrine Metabolic Ovarian Syndrome (PMOS),^12^ with hyperandrogenism (i.e. elevated testosterone levels) as a key diagnostic feature,^13^ is associated with 68% higher odds of CVD according to a meta-analysis based on 1.06 million women^14^ with insulin resistance (IR) as a key pathophysiological feature.^15^

Dietary interventions, particularly dietary fibre supplementation, are safe and cost-effective strategies for CVD prevention.^16–18^ Fibre prevents CVD via several mechanisms. For example, fibre lowers LDL cholesterol^19^ and blood pressure (BP),^17,18,20^ both key risk factors for CVD.^18^ Hypertension, in particular, is a more significant CVD risk factor for females compared to males.^6,21^ Short-chain fatty acids (SCFAs, primarily acetate, propionate, and butyrate), key metabolites produced through gut microbiota fermentation of fibre,^22^ are essential for how fibre prevents CVD.^16,23^ SCFAs reduce CVD risk by suppressing inflammatory pathways in immune cells via activation of G protein-coupled receptors (e.g., GPR41, GPR43, GPR65, GPR68).^24–28^ Moreover, SCFAs improve cardiometabolic health by stimulating glucagon-like peptide-1 (GLP-1) secretion,^29^ lowering blood glucose and appetite.^30^ A remaining question is whether SCFAs can reduce the excessive CVD risk in high-risk females.

Therefore, we employed acetate, the most abundant SCFA that reaches systemic circulation, as a surrogate marker for dietary fibre intake in two large, ethnically diverse cohorts with long-term CVD outcomes. We aimed to examine whether circulating acetate modifies the association between MACE risk and altered sex hormonal profiles (i.e., early menopause, representing estrogen deficiency, and elevated free testosterone (FT), a key diagnostic feature of PMOS^31^). We hypothesised that high acetate levels would be associated with reduced MACE incidence in high-risk females.

## Methods

### UK Biobank Data Extraction

The UK Biobank (approval 86879) is a comprehensive prospective cohort study involving approximately 500,000 participants recruited from 2006 to 2011 across the United Kingdom.^32^ At the initial visit at the assessment centre, participants provided demographic information, including age and body mass index (BMI), and health-related data such as medication use and age at menopause (Tables 1 and S1). Blood pressure (BP) was measured twice, within a few minutes, using automated sphygmomanometers. Blood and urine samples were also collected during this initial visit. Plasma metabolites, including acetate, triglycerides, and cholesterol, were quantified by nuclear magnetic resonance (NMR) spectroscopy at Nightingale Health. Plasma total testosterone and sex hormone-binding globulin (SHBG) were measured using the UniCel DxI 800 Access Immunoassay System. Spot urinary potassium and sodium levels were assessed using photometry. Information on hormone replacement therapy (HRT), a commonly used medication to reduce menopausal symptoms, was obtained from questionnaires (Table S1).

Major adverse cardiovascular events (MACE) were used as the primary outcome to evaluate cardiovascular health (Figure S1, Table S1). MACE was identified using hospital admissions, surgical procedures, and deaths attributed to acute coronary syndrome (ACS), heart failure (HF), and ischemic stroke, as previously reported^26,33^. Plasma acetate measured by NMR was used as a surrogate for dietary fibre intake, as acetate accounts for over 60% of SCFAs by molar proportion in gut fibre fermentation.^34^ Since fibre and alcohol are the major sources of acetate, linear models without the intercept were constructed to assess the association between fibre intake from a 24h dietary recall and plasma acetate, with alcohol intake, standardised age and body mass index (BMI) adjusted. Spot urinary sodium and potassium levels were incorporated as precise measures of dietary sodium and potassium intake, respectively,^35^ important dietary factors influencing cardiovascular health.^36^ Participants who did not report current HRT use but had a record of HRT were categorised as previous HRT users.

Data from female participants with complete medical records, BP, plasma acetate, alcohol intake frequency, urinary potassium and sodium, BMI, HRT record, and medication intake information were extracted. Participants taking insulin or contraceptive pills were excluded, as these may affect the endogenous production of sex hormones.^37^ Participants taking cholesterol-lowering or blood pressure medication were classified as taking CVD-preventive medications. We excluded participants younger than 45 years due to a lack of information on menopause after recruitment and those with any pre-existing MACE before recruitment (Figure S1). Participants without complete menopausal status reporting or having a hysterectomy, and participants whose reported menopause age did not align with their menopause status, as well as those reporting menopause after age 60,^38,39^ were excluded.

### Early menopause

Early menopause was defined as menopause before age 45 (Figure S1). We used ‘early menopause’ as a surrogate for low oestradiol levels due to: (i) the limited sample size (∼76k out of ∼500k participants) of participants with measured oestradiol levels, and (ii) a competitive assay was used to measure oestradiol levels, which is less accurate than mass spectrometry, especially at low concentrations.^40^ Therefore, postmenopausal participants were required to report their age at menopause; those who did not were excluded. Participants with a history of bilateral oophorectomy who did not report the timing of the procedure were also excluded. To ensure accurate reporting, individuals who underwent bilateral oophorectomy but did not report menopause in the same year were excluded. Premenopausal participants or those without a bilateral oophorectomy history were retained in the whole cohort.

### Biobank Japan Data Analysis

We used Biobank Japan (BBJ) to validate findings regarding early menopause. BBJ is a prospective hospital-based biobank ^41–43^ with plasma metabolite levels measured by NMR (Nightingale Health) available for 105,494 participants, with 23,764 older than 45 years being eligible for this study (Table 2). The same outcomes as per the UKBB were assessed. In addition to standard internal quality control, technical variation was corrected using the R package *ukbnmr.*^44^ Participants younger than 45 or older than 85 years, those with values more than 3 times the interquartile range (IQR) or 3 standard deviations (SD) from the mean of acetate, or pre-existing CVD were also excluded. Moreover, only participants with ≥1 year of follow-up were included in the time-to-event analysis of MACE.

### Analysis of free testosterone

PMOS is one of the most common diseases in females caused by dysregulation of sex hormones. However, because the number of UK Biobank participants with diagnosed PMOS is limited (only 444 cases in the whole database), we used FT as a surrogate for PMOS.^31,45^ The free testosterone (FT) level was calculated using the Vermeulen equation,^46^ based on the total testosterone (Total T) level measured by an immunoassay using the formula:

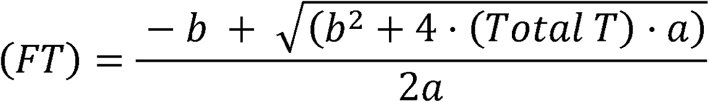

Where:

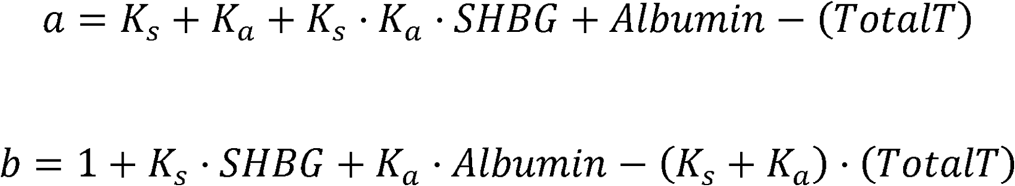

SHBG stands for the concentration of sex hormone-binding globulin. *K_S_* = 1×109 *L*/*mol* and *K_a_* = 3.6×104 *L*/*mol* are the association constants of testosterone with SHBG and albumin, respectively.

We defined elevated FT as a plasma FT level above 31.2 pmol/L (9 pg/mL).^47,48^ Because high FT levels in females have been associated with insulin resistance,^15^ we also included the triglyceride to high-density lipoprotein cholesterol (TG/HDL-C) ratio, a surrogate marker of insulin resistance, as a covariate.^49,50^

### Plasma proteomics analysis

To explore underlying mechanisms, we examined plasma proteins associated with early menopause, elevated FT, and acetate. Proteomic profiling was conducted on ∼50,000 plasma samples from UK Biobank participants using the antibody-based Olink Explore 3072 PEA platform, yielding plasma levels of 2,941 proteins^51^. Participants with an analytic sample and valid paired plasma proteomics data were included in the proteomics analysis. Proteins detected in fewer than 50% of included samples or in samples with fewer than 20% of proteins detected were considered low-quality and excluded, leaving 9,018 participants with 2,920 high-quality plasma proteins for downstream analysis. Upregulated proteins were defined as log_2_(fold change) > 0.05 and false discovery rate (FDR) < 0.05, and downregulated proteins were defined as log_2_(fold change) <0.05 and FDR < 0.05 and calculated using limma (3.60.4)^52^, with alcohol consumption, standardised age and BMI, urine potassium and sodium, current and previous HRT use, and also CVD-preventive medication intake adjusted. We analysed whether pro-inflammatory and pro-fibrotic pathways were enriched among proteins positively correlated with early menopause and negatively correlated with acetate. Pathway enrichment analysis was performed using clusterProfile^53^ using all 2,920 detected plasma proteins as background.

### Statistics

We first assessed the associations between 10-year MACE, plasma acetate, early menopause, and elevated FT in the UK Biobank. Subsequently, to assess whether plasma acetate modifies the cardiovascular risk associated with early menopause or elevated FT, we compared the 10-year MACE incidence associated with these conditions between participants with plasma acetate levels above and below the median. We also conducted additional stratified analyses across acetate quartiles.

The hazard ratio (HR) of early menopause or elevated FT for MACE by acetate level was further assessed by stratifying the dataset into acetate percentiles. The HR curves by percentile were smoothed using a generalised linear model (GLM) or local regression (LOESS). Finally, we examined the interaction between standardised plasma acetate level and early menopause or elevated FT. Interaction terms with other covariates were also inserted into the model to avoid spurious non-linear effects.^54^ As a sensitivity analysis, the stratification, interaction, and percentile analyses were repeated after restricting the sample to postmenopausal participants (“Postmenopausal Cohort”).

All time-to-event models in the UK Biobank were adjusted for age, BMI, CVD-preventive medication, alcohol intake, and urine sodium and potassium. Moreover, in the early menopause analysis, current or previous HRT use was also adjusted. For free testosterone, the TG/HDL-C ratio was also adjusted as the surrogate for insulin resistance. Time-to-event analyses were performed using Cox proportional hazards models, implemented with the R packages survival (version 3.6.4) and survminer (version 0.4.9).^55^ *P*<0.05 was considered significant.

## Results

### Baseline characteristics of the UK Biobank participants

After applying the exclusion criteria, a total of 81,799 participants (Table 1, Figure S1) were retained for the early menopause analyses. As part of the sensitivity analysis, we additionally extracted a subset of postmenopausal participants (n=68,249) for early-menopause analyses, hereafter referred to as the “postmenopausal cohort” (Table 1, Figure S1). For the analysis of FT, participants without valid TG/HDL-c ratios or calculated FT values were excluded, yielding 59,247 participants.

**Table 1.** Demographics of the UK Biobank participants included for early menopause analysis.

| Whole Cohort (n=81,799) |  |  |  |  |
| --- | --- | --- | --- | --- |
|  | All included participants<br>(n=81,799) | Early menopause<br>(n=8,840) | Others<br>(n=72,959) | P-value |
| Age, yr | 59 (53-63) | 61 (55-64) | 59.0 (53-63) | <0.001 |
| BMI, kg/m<sup>2</sup> | 26.9 ± 4.9 | 27.4 ± 5.1 | 26.9 ± 4.9 | <0.001 |
| SBP, mmHg | 136.9 ± 19.1 | 137.7 ± 19.4 | 136.8 ± 19.1 | <0.001 |
| DBP, mmHg | 80.8 ± 9.9 | 80.8 ± 9.9 | 80.8 ± 9.9 | 0.8 |
| Plasma acetate, µmol/L | 15.7 ± 5.8 | 15.3 ± 5.8 | 15.7 ± 5.8 | <0.001 |
| Current HRT use | 4,582 (5.6%) | 963 (10.9%) | 3,619 (5.0%) | <0.001 |
| Previous HRT use | 27,290 (33.4%) | 4,658 (52.7%) | 22,632 (31.0%) | <0.001 |
| CVD-preventive medication intake | 19,163 (23.4%) | 2,611 (29.5%) | 16,552 (22.7%) | <0.001 |
| Regular drinker | 62,688 (76.6%) | 6,297 (71.2%) | 56,391 (77.3%) | <0.001 |
| Urinary sodium, mmol/L | 65.3 ± 38.8 | 66.6 ± 39.2 | 65.2 ± 38.7 | <0.001 |
| Urinary potassium, mmol/L | 58.5 ± 32.7 | 59.3 ± 33.0 | 58.4 ± 32.7 | 0.011 |
| Postmenopausal Cohort (n=68,249) |  |  |  |  |
|  | All included participants<br>(n=68,249) | Early menopause<br>(n=8,840) | Others<br>(n=59,409) | P-value |
| Age, yr | 61 (56 -64) | 61 (55-64) | 61 (56-64) | <0.001 |
| BMI, kg/m<sup>2</sup> | 27.0 ± 4.9 | 27.4 ± 5.1 | 26.9 ± 4.9 | <0.001 |
| SBP, mmHg | 138.4 ± 19.1 | 137.7 ± 19.4 | 138.5 ± 19.1 | <0.001 |
| DBP, mmHg | 81.0 ± 9.8 | 80.8 ± 9.9 | 81.0 ± 9.8 | 0.011 |
| Plasma acetate, µmol/L | 15.7 ± 5.9 | 15.3 ± 5.8 | 15.8 ± 5.9 | <0.001 |
| Current HRT use | 4,165 (6.1%) | 963 (10.9%) | 3,202 (5.4%) | <0.001 |
| Previous HRT use | 26,893 (39.4%) | 4,658 (52.7%) | 22,235 (37.4%) | <0.001 |
| CVD-preventive medication intake | 17,940 (26.3%) | 2,611 (29.5%) | 15,329 (25.8%) | <0.001 |
| Regular drinker | 51,802 (75.9%) | 6,297 (71.2%) | 45,505 (76.6%) | <0.001 |
| Urinary sodium, mmol/L | 64.0 ± 37.7 | 66.6 ± 39.2 | 63.7 ± 37.5 | <0.001 |
| Urinary potassium, mmol/L | 58.6 ± 32.5 | 59.3 ± 33.0 | 58.5 ± 32.4 | 0.035 |

**Legend:** BMI, body-mass index; SBP, systolic blood pressure. DBP, diastolic blood pressure. HRT, hormone replacement treatment. Continuous data were presented as median (quartiles 1- 3) or mean ± standard deviation. Categorical data were presented as counts (percentages). The p-values comparing participants with early menopause and other participants were calculated using Wilcoxon-Mann-Whitney for continuous data, and $\chi^2$ test for categorical data)

### Plasma acetate and long-term cardiovascular risk in the whole cohort

Each 1g/day increase in fibre intake was associated with 0.505 μmol/L higher plasma acetate after adjusting for alcohol intake and standardised age and BMI, according to a regression model without an intercept (adjusted R^2^ = 0.813; Table S2). In the subset of non-regular drinkers, each 1g/day increase in fibre intake was associated with 0.706 μmol/L higher plasma acetate with standardised age and BMI adjusted (adjusted R^2^ = 0.740; Table S2). Participants with higher plasma acetate levels had a significantly lower incidence of MACE within 10 years of recruitment (Figure 1A). Acetate levels above the median (15.2 μmol/L, equal to ∼21.6 g/day fibre intake in non-regular drinkers with average age and BMI) were associated with a lower 10-year incidence of MACE (−6.3/1000 woman-year, HR=0.887, 95% CI:[0.823-0.955], *p*=0.002; Figure 1B). Similarly, acetate levels above the median group were significantly associated with a lower 10-year incidence of MACE in both the postmenopausal cohort (Figure 1C) and non-regular drinkers (Figure S2A). In another model treating acetate as a continuous variable with plasma acetate × covariates interaction terms included (Table S3), a 1-standard deviation (SD) increase of acetate was significantly associated with lower MACE incidence (Whole: HR = 0.885, 95% CI:[0.813-0.964], *p*=0.005; Postmenopausal: HR = 0.867, 95% CI:[0.791-0.951], *p*=0.002) in participants with average BMI, age, and urine sodium and potassium levels not taking CVD-related medication or HRT. Similarly, in analogous linear models with acetate × covariates interaction terms (Table S4), a 1-SD increase in plasma acetate was associated with - 0.373 mmHg lower SBP in participants with mean BMI, age, urine sodium and potassium, and not taking CVD-related medication or HRT.

**Figure 1.**
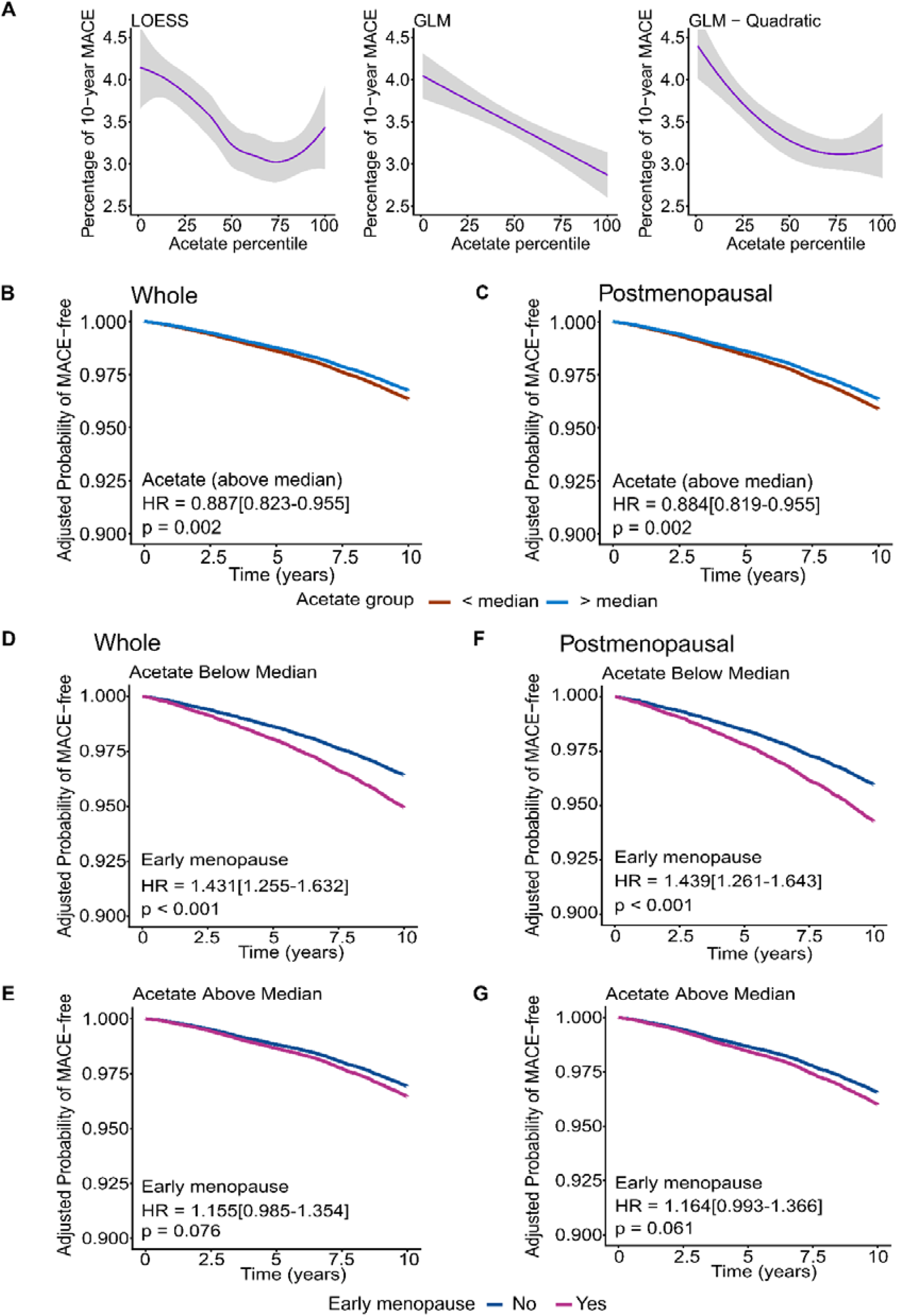
Association of plasma acetate and early menopause on 10-year incidence of major adverse cardiovascular events (MACE) in UK Biobank. **A.** Comparison of percentage of participants experiencing MACE within 10 years of recruitment according to acetate percentile in the whole cohort (n=81,799). Smoothed curves were derived from locally estimated scatterplot smoothing (LOESS), generalised linear regression model (GLM) and GLM including a quadratic term for the adjusted coefficient size. **B-C.** Time-to-event analysis comparing adjusted 10-year MACE incidence between participants with acetate above versus below the median in the UK Biobank in **B.** the whole cohort (n=81,799) and **C.** the postmenopausal cohort (n=68,249). **D-G.** Time-to-event analysis of MACE incidence associated with early menopause stratified by plasma acetate level (above vs. below median) in UK Biobank for 10 years in **D-E.** the whole cohort (n=81,799) and **F-G.** the postmenopausal cohort (n=68,249). All analyses were adjusted for alcohol consumption, standardised age and BMI, urine potassium and sodium, current and previous HRT use, and also CVD-preventive medication intake.

### Early menopause, acetate, and long-term cardiovascular risk

Early menopause participants showed higher BMI, age, and a greater proportion of current or previous hormone replacement treatment (HRT) use and CVD-related medication intake (Table 1). Early menopause was associated with increased MACE incidence in both the whole cohort (Figure S2B; HR=1.310, 95% CI: [1.184-1.450], *p*<0.001) and postmenopausal cohort (Figure S2C; HR=1.320, 95% CI: [1.192-1.461], *p*<0.001) with age, BMI, urinary potassium and sodium, alcohol consumption, CVD-preventive medicine intake and current or previous HRT use adjusted. After grouping participants by plasma acetate levels above or below the median, among those with levels below the median, early menopause was associated with a higher incidence of MACE (Figure 1D; HR=1.431, 95% CI: [1.255-1.632], *p*<0.001). No significant difference in MACE incidence was observed between early- and non-early-menopause participants when plasma acetate levels were above the median (Figure 1E; HR=1.155, 95% CI: [0.985-1.354], *p*=0.075). The sensitivity analyses, including only postmenopausal participants and non-regular drinkers, showed similar results (Figures 1F-G; S2D-E).

### Baseline characteristics of the Biobank Japan participants and validation

After filtering for the same selection criteria used in the UK Biobank, 23,764 participants from the Biobank Japan were included in the replication analysis (median follow-up year=6.9; maximum follow-up year=9.7, Table 2). Among participants with plasma acetate levels below the median in Biobank Japan, early menopause was significantly associated with a higher incidence of MACE (Figure 2A; HR=1.423, 95% CI: [1.040-1.948], *p*=0.028). Conversely, among participants with plasma acetate levels above the median, no significant association was observed between early menopause and MACE incidence (Figure 2B; HR=1.350, 95% CI: [0.979-1.862], *p*=0.067). Similarly, in the sensitivity analysis including only post-menopausal participants (n=19,031), the significant association between early menopause and MACE incidence was observed only among participants with plasma acetate levels below the median (Figures 2C-D).

**Figure 2.**
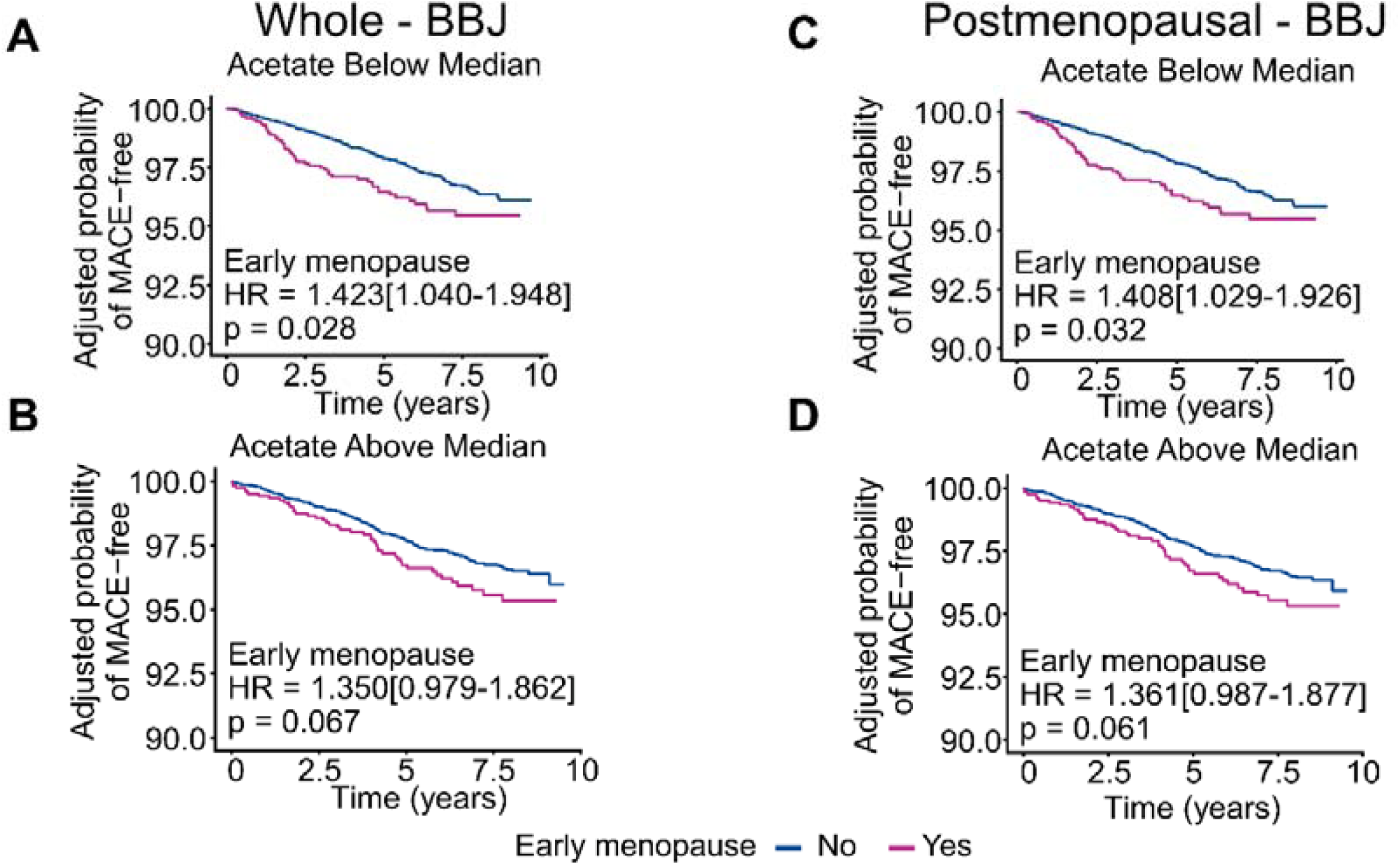
Association of plasma acetate and early menopause with the incidence of major adverse cardiovascular events (MACE) in Biobank Japan. Time-to-event analysis of MACE incidence associated with early menopause stratified by plasma acetate level (above vs. below median) in Biobank Japan in **A-B.** the whole cohort (n=23,764), and **C-D.** the postmenopausal cohort (n=19,031); median follow-up year = 6.9; maximal follow-up years=9.7. All analyses were adjusted for age and BMI.

**Table 2.** Demographics of the Biobank Japan participants included.

| Whole Cohort (n=23,764) |  |  |  |  |
| --- | --- | --- | --- | --- |
|  | All included participants<br>(n=23,764) | Early menopause<br>(n=2,384) | Others<br>(n=21,380) | P-value |
| Age, yr | 65 (57-72) | 66.0 (58-72) | 65 (57-72) | <0.001 |
| BMI, kg/m² | 23.1 ± 3.6 | 23.2 ± 3.7 | 23.1 ± 3.6 | 0.034 |
| SBP, mmHg | 130.2 ± 16.8 | 130.6 ± 17.2 | 130.2 ± 16.8 | 0.25 |
| DBP, mmHg | 75.8 ± 10.6 | 76.0 ± 10.4 | 75.8 ± 10.6 | 0.47 |
| Plasma acetate, µmol/L | 33.4 ± 19.0 | 33.9 ± 19.2 | 33.3 ± 18.9 | 0.17 |
| CVD-preventive medication intake | 7954 (33.5%) | 865 (36.3%) | 7089 (33.2%) | 0.0023 |
| Regular drinker | 5166 (22.2%) | 470 (20.1%) | 4696 (22.5%) | 0.011 |
| Postmenopausal Cohort (n=19,031) |  |  |  |  |
|  | All included participants<br>(n=19,031) | Early menopause<br>(n=2,384) | Others<br>(n=16,647) | P-value<br>(EM vs others) |
| Age, yr | 66 (59-72.0) | 66 (58-72) | 66 (59-72) | 0.042 |
| BMI, kg/m² | 23.1 ± 3.6 | 23.2 ± 3.7 | 23.1 ± 3.6 | 0.041 |
| SBP, mmHg | 130.8 ± 16.8 | 130.6 ± 17.2 | 130.8 ± 16.7 | 0.55 |
| DBP, mmHg | 75.9 ± 10.5 | 76.0 ± 10.4 | 75.9 ± 10.6 | 0.57 |
| Plasma acetate, µmol/L | 33.7 ± 19.2 | 33.9 ± 19.2 | 33.7 ± 19.2 | 0.78 |
| CVD-preventive medication intake | 6748 (35.5%) | 865 (36.3%) | 5883 (35.3%) | 0.38 |
| Regular drinker | 3888 (20.9%) | 470 (20.1%) | 3418 (21.0%) | 0.37 |

**Legend:** BMI, body-mass index; EM, early menopause; sSBP, systolic blood pressure. DBP, diastolic blood pressure. Continuous data were presented as median (quartiles 1- 3) or mean ± standard deviation. Categorical data were presented as counts (percentages). The p-values comparing participants with early menopause and other participants were calculated using Wilcoxon-Mann-Whitney for continuous data, and $\chi^2$ test for categorical data.

### Early menopause × acetate interaction on long-term cardiovascular risk

Analysis of the HR for MACE associated with early menopause across acetate percentiles showed that the HR was lower in participants with higher acetate levels in both the whole and postmenopausal cohorts (Figures 3A-B). Subsequently, we performed a stratification analysis by acetate quartile (Quartile 1 (Q1): < 16.7g/fibre/day; Q2: 16.7 – 21.6g/fibre/day; Q3: 21.6 – 27.1g/fibre/day; Q4: > 27.1g/fibre/day). Consistent with the percentile-based analysis, the association between early menopause and MACE was not significant among participants with acetate levels in Q3 or Q4 (Figures S3A-B) in either cohort. Similarly, in non-regular drinkers, the association between early menopause and MACE was only significant in participants with acetate levels in Q1 (HR=1.416, 95% CI: [1.053-1.905], *p*=0.021, Figure S3C). Furthermore, acetate was modelled as a continuous variable in a model with both acetate × covariate and early menopause × covariate interactions. The acetate × early menopause interaction was significantly associated with a lower incidence of MACE in both cohorts (Figure 3C; Table S5; Whole Cohort: HR=0.894; 95% CI: [0.804-0.993]; *p*=0.037; Postmenopausal Cohort: HR=0.898; 95% CI: [0.808-0.998]; *p*=0.046).

**Figure 3.**
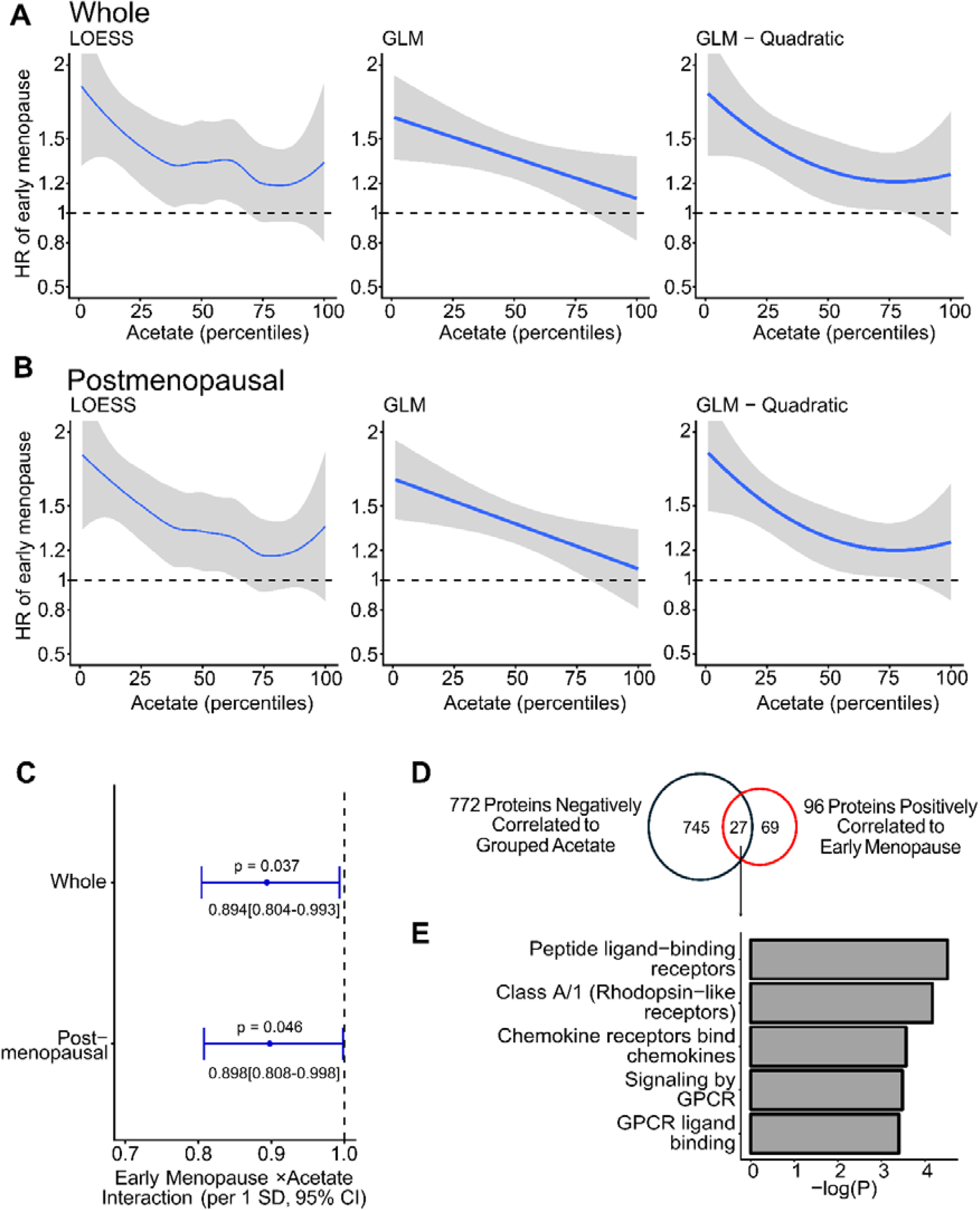
Plasma acetate modifies the association between early menopause on 10-year incidence of major adverse cardiovascular events (MACE) in UK Biobank. **A-B.** Smoothed curves derived from locally estimated scatterplot smoothing (LOESS), generalised linear regression model (GLM) and GLM including a quadratic term for the adjusted coefficient size of early menopause across different percentiles of plasma acetate levels in **A.** the whole cohort (n=81,799) and **B.** the postmenopausal cohort (n=68,249) on the MACE hazard ratio. **C.** Coefficient sizes and 95% confidence intervals (CIs) (shown as error bars) for the interaction term between plasma acetate and early menopause across the whole and the postmenopausal cohort (n=81,799 and 68,249) on 10-year MACE incidence. **D.** Venn diagram showing the overlap between overrepresented plasma proteins in participants with early menopause and underrepresented plasma proteins in participants with an acetate level above the median. **E.** Pathway enrichment analysis (Reactome database) of proteins that are both overrepresented plasma proteins in participants with early menopause and underrepresented plasma proteins in participants with an acetate level above the median in the UK Biobank. All analyses were adjusted for alcohol consumption, standardised age and BMI, urine potassium and sodium, current and previous HRT use, and also CVD-preventive medication intake (n=9,018).

### Free testosterone, acetate, and long-term cardiovascular risk

Elevated FT was analysed in 59,247 participants from the UK Biobank (Table S6). Elevated FT was also associated with higher MACE incidence (Figure S4A; HR= 1.206, 95% CI: [1.026-1.419], *p*=0.024). Among participants with plasma acetate levels below the median, elevated FT was associated with a higher incidence of MACE (Figure S4B; HR= 1.283, 95% CI: [1.047-1.573], *p*=0.016). Similarly to early menopause, no significant difference in MACE incidence was observed between elevated and unelevated FT participants in the group with plasma acetate levels above the median (Figure S4C; HR=1.080, 95% CI: [0.825-1.415], *p*=0.576).

Analysis of the HR for MACE associated with elevated FT across acetate percentiles showed that the HR for MACE was lower in participants with higher acetate levels (Figure S4D). Nevertheless, the acetate × elevated FT interaction was also not significantly associated with MACE incidence (Table S7, *p*=0.24) in the model with both acetate × covariate and elevated FT × covariate interactions. Similarly, in an analogous model that treated FT as a continuous variable, the acetate × FT interaction was not significantly associated with MACE incidence (Table S7, *p*=0.09).

### Early menopause, elevated FT, acetate, and plasma proteomics

Plasma proteomics identified 772 proteins underrepresented in participants with acetate levels above the median; these were predominantly pro-inflammatory on pathway enrichment analysis (Tables S8-S9). In contrast, 51 proteins were overrepresented with higher acetate, with significant metabolic pathway enrichment (Tables S8-S9). Early menopause was associated with 96 overrepresented and 21 underrepresented proteins, with the overrepresented proteins showing enrichment for pro-inflammatory pathways (Tables S8 and S10). Elevated FT was associated with 152 overrepresented and 49 underrepresented proteins, and the overrepresented proteins were predominantly associated with oxidative stress (Tables S8 and S11). Notably, 27 pro-inflammatory proteins overlapped between proteins overrepresented in early menopause and those underrepresented with higher acetate (Figures 3D-E, Tables S8 and S12). This included CCL17 and CCL22. No protein was both underrepresented in early menopause and overrepresented at higher acetate levels. Conversely, 49 pro-fibrotic proteins overlapped between proteins overrepresented in elevated FT and those underrepresented with higher acetate (Tables S8 and S12). These proteins included MMP8, MMP9, and COL2A1. Only 6 proteins were both underrepresented in participants with elevated FT and overrepresented in participants with higher acetate (Table S12), including IGFBP1 and IGFBP2.

## Discussion

This study investigated the impact of plasma acetate, the most abundant SCFA produced by the gut microbiota via fibre fermentation, on cardiovascular health in females. Previous studies with smaller sample sizes (n=233 to 3,129) have reported inconsistent findings regarding the association between plasma acetate levels and CVD risk.^56–59^ In contrast, our study, conducted in a substantially larger cohort (n=81,799), demonstrated that higher plasma acetate levels, particularly equivalent to >27.1g/fibre/day (Q4), were associated with a lower risk of MACE and lower BP in females. Our findings are impactful, as the ∼10% lower hazard of CVD associated with higher acetate levels is comparable to the risk reductions reported for population level sodium reduction strategies.^60^ We then examined high-risk groups where sex hormone levels are altered. We demonstrated that higher plasma acetate levels were associated with a lower 10-year incidence of CVD events in early menopause, as validated by two independent cohort studies with different ancestries, the UK Biobank and Biobank Japan. Using plasma proteomics, we identified that this may be via a decrease in pro-inflammatory mechanisms. Together, these findings highlight the potential of dietary fibre or acetate supplementation as a cost-effective and readily available therapy, offering a promising avenue for reducing cardiovascular risk in females, particularly those with early menopause.

Early menopause is characterised by abnormally low oestrogen in females.^9,11^ Our findings of higher MACE incidence in females with early menopause are consistent with previous studies in European populations.^61^ Moreover, this association is also relevant for individuals with East Asian ancestry. Notably, we found that higher plasma acetate levels were associated with lower cardiovascular risk in those with early menopause, which was further supported by a significant negative interaction term between acetate and early menopause on MACE incidence. The maximum benefit observed in the top quartile is equivalent to 27.1g/fibre/day, which is near the threshold recommended for females with hypertension;^62^ however, this recommendation is lacking in guidelines specific to early menopause.

Early menopause is associated with higher inflammation and oxidative stress.^63^ In contrast, the cardioprotective effects of SCFAs are largely mediated through anti-inflammatory signalling via G protein-coupled receptors GPR41, GPR43, and GPR65.^20,25,26,64^ Indeed, our findings indicate that pro-inflammatory plasma proteins associated with early menopause were negatively correlated with plasma acetate, supporting the hypothesis that inflammation mediates the association between early menopause and acetate in cardiovascular health. These findings suggest that early menopause may involve an endogenous pro-inflammatory transition, which could be counteracted by the anti-inflammatory properties of acetate via activation of GPR41/43 or GPR65.^20,27^

Elevated testosterone is also a significant risk factor for CVD in females. Testosterone plays markedly different roles in males and females, reflecting distinct mechanisms of action.^65,66^ While there is still no consensus on whether testosterone in men is associated with cardiovascular risk,^67,68^ elevated testosterone levels have been implicated not only in reproductive disorders such as PMOS,^69,70^ but also in higher cardiovascular risk in women.^71^

Testosterone has been associated with the activation of pro-inflammatory and pro-fibrotic signalling pathways,^70,72,73^ which might contribute to long-term cardiovascular risk. Indeed, we found pro-fibrotic proteins both overrepresented in elevated FT and underrepresented with higher acetate. In stratification analyses, we found that elevated FT levels were associated with a higher incidence of MACE, and this risk might be mitigated by higher plasma acetate levels.

Nevertheless, the interaction term between acetate and elevated FT levels was non-significant. These results suggest that further study with a larger sample size is warranted to clarify the potential interplay between acetate and elevated FT on cardiovascular risk. Although the modification of acetate in the association of elevated FT with cardiovascular risk is only suggestive, the overlap in the associated proteomic signature supports the notion that higher plasma acetate may counteract the pro-fibrotic signals of elevated FT in women.

We acknowledge that this study has several limitations. Firstly, while our findings support an interaction between early menopause, acetate, and cardiovascular risk, causality cannot be inferred from the current data. Future clinical trials are warranted to validate the findings, and animal model studies are warranted to confirm the underlying mechanisms. Secondly, to our knowledge, the UK Biobank is the only large-scale cohort dataset that simultaneously includes plasma acetate, testosterone, and CVD data. Because Biobank Japan lacked plasma testosterone data, validations regarding FT could not be conducted. Moreover, plasma testosterone levels in the UK Biobank were measured by immunoassay rather than mass spectrometry, which is considered the gold standard. Finally, the lack of plasma propionate and butyrate data in both the UK Biobank and Biobank Japan precludes investigation of the associations between these two less abundant SCFAs and cardiovascular outcomes.

In conclusion, higher plasma acetate levels were associated with lower cardiovascular risk in females, with the greatest benefit observed in those with early menopause. These findings suggest that both microbial acetate and its dietary source – fibre – may mitigate hormone related cardiovascular vulnerability, underscoring the importance of hormonal context when developing prevention strategies to reduce CVD in females.

## Acknowledgement

This research has been conducted using the UK Biobank Resource under Application Number 86879. Monash eResearch, including the M3 servers, supported this research.

## Author Contributions

Concept and design: CY, LM, AV, FZM. Acquisition, analysis and interpretation of data: CY, SN, KM, YO, NT, KL, LM, AV, FZM. Drafting of manuscript: CY, SN, KM, YO, FZM. Critical revision of manuscript: LM, AV. Statistical analysis: CY. Obtained funding, access to UKBB, and supervision: FZM. All authors read and approved the manuscript.

## Data availability

The R codes are available at https://github.com/ChrYang/SexHormone.

