## Supplementary material for "Acetate, a fibre-derived gut metabolite, is associated with reduced cardiovascular disease risk in females with early menopause": Online supplementary figures

**Running title:** Dietary fibre, sex hormones and cardiovascular disease

Chaoran Yang^1,2^, Shinichi Namba^3,4,5^, Koichi Matsuda^6,7^, Yukinori Okada^3,4,5,8,9^, BioBank Japan Project, Nele Taba^10^, Kreete Lüll^10^, Lisa Moran^2,11#^, Amanda Vincent^11#^, Francine Z. Marques^1,2,12^*

^1^Hypertension Research Laboratory, Department of Pharmacology, Biomedical Discovery Institute, Faculty of Medicine, Nursing and Health Sciences, Monash University, Melbourne, Australia; ^2^Victorian Heart Institute, Monash University, Melbourne, Australia; ^3^Department of Genome Informatics, Graduate School of Medicine, The University of Tokyo, Tokyo, Japan; ^4^Department of Statistical Genetics, Osaka University Graduate School of Medicine, Suita, Japan; ^5^Laboratory for Systems Genetics, RIKEN Center for Integrative Medical Sciences, Yokohama, Japan; ^6^Laboratory of Clinical Genome Sequencing, Department of Computational Biology and Medical Sciences, Graduate School of Frontier Sciences, The University of Tokyo, Tokyo, Japan; ^7^Laboratory of Genome Technology, Human Genome Center, Institute of Medical Science, The University of Tokyo, Tokyo, Japan; ^8^Laboratory of Statistical Immunology, Immunology Frontier Research Center (WPI-IFReC), Osaka University, Suita, Japan; ^9^Premium Research Institute for Human Metaverse Medicine (WPI-PRIMe), Osaka University, Suita, Japan; ^10^Institute of Genomics, Faculty of Science and Technology, University of Tartu, Estonia; ^11^Monash Centre for Health Research and Implementation (MCHRI), Monash University, Melbourne, Australia; ^12^Baker Heart and Diabetes Institute, Melbourne, Australia

\# contributed equally

***Corresponding author**: Prof Francine Marques, Hypertension Research Laboratory, Victorian Heart Institute, Level 2, Victorian Heart Hospital, 631 Blackburn Road Clayton, VIC 3168 Monash University, Melbourne, Australia, Phone: +61-03-9905 6958.

**Supplemental Figures**

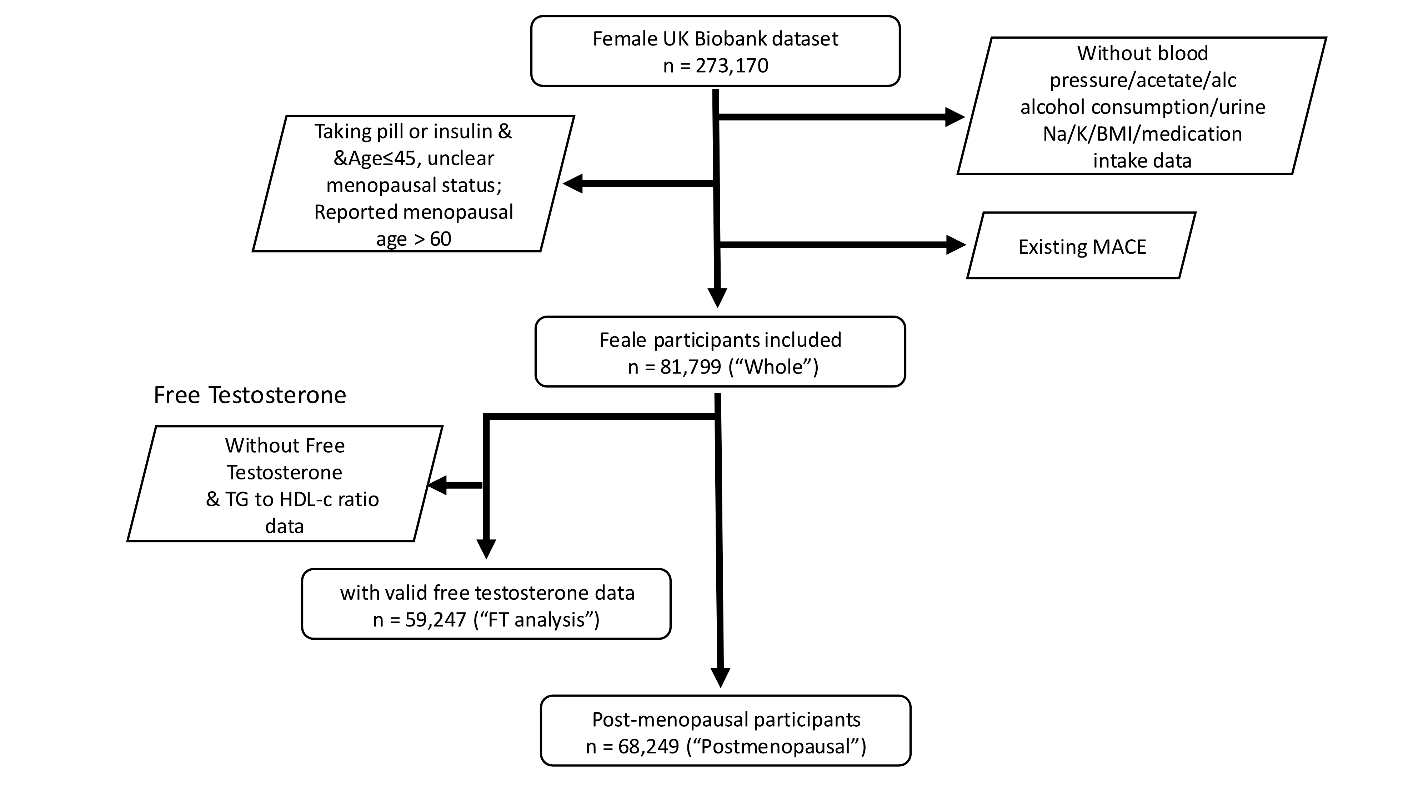

**Figure S1** | **Data inclusion criteria**. Flowchart of the data filtering pipeline.

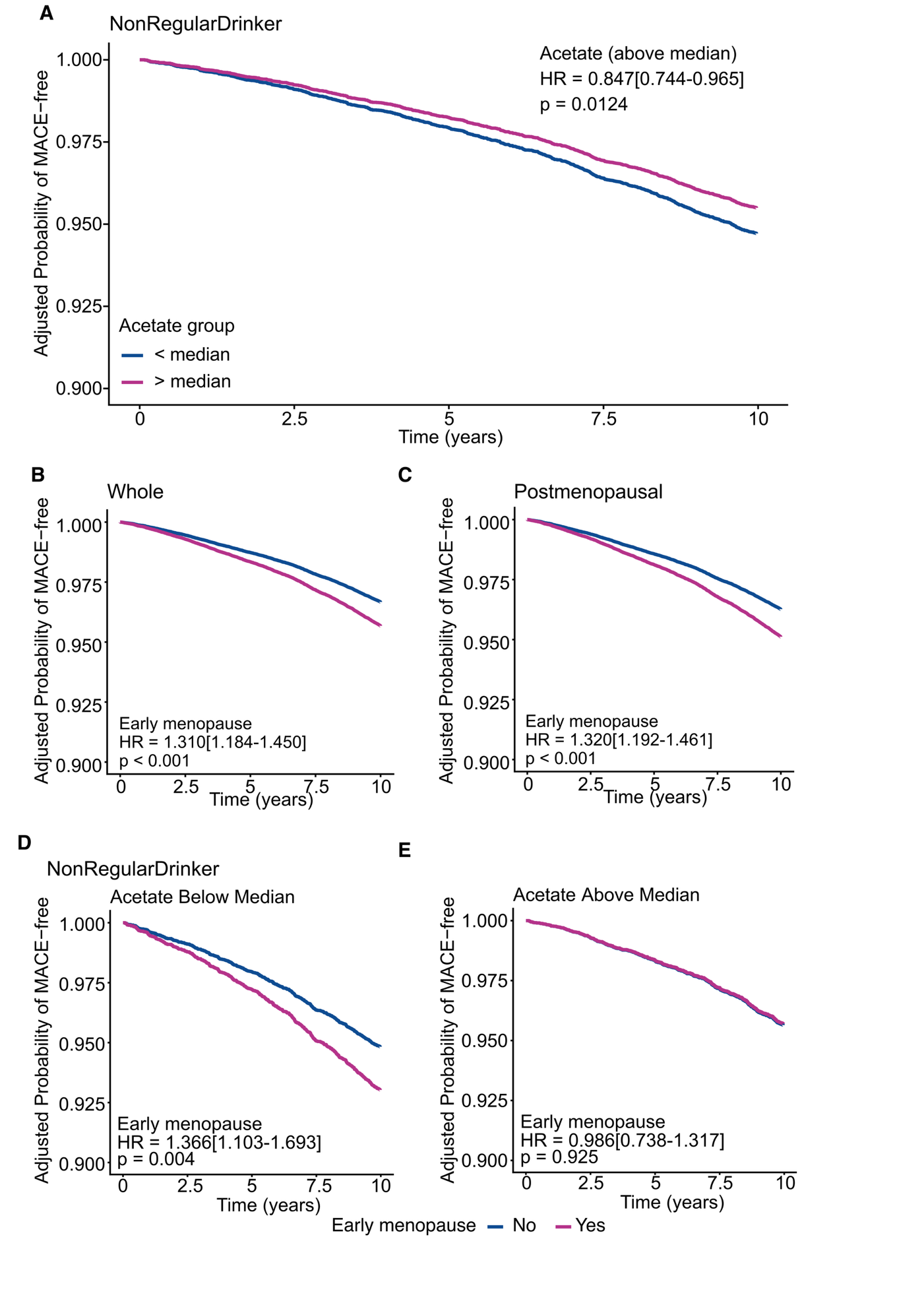

**Figure S2** | **Association of plasma acetate and early menopause on 10-year incidence of major adverse cardiovascular events (MACE) in UK Biobank. A.** Time-to-event analysis comparing adjusted 10-year MACE incidence between participants with acetate above versus below the median in the UK Biobank in non-regular drinkers. (N=19,111) **B-C.** Time-to-event analysis of adjusted 10-year MACE incidence associated with early menopause in **B.** the whole cohort (N=81,799) and **C.** the postmenopausal cohort (N=68,249). **D-E.** Time-to-event analysis of MACE incidence in early menopause versus non-early menopause participants stratified by plasma acetate level (above vs. below median) in UK Biobank for 10 years in non-regular drinkers. (N=19,111). All analyses were adjusted for alcohol consumption, standardised age and BMI, urine potassium and sodium, current and previous HRT use, and also CVD medication intake.

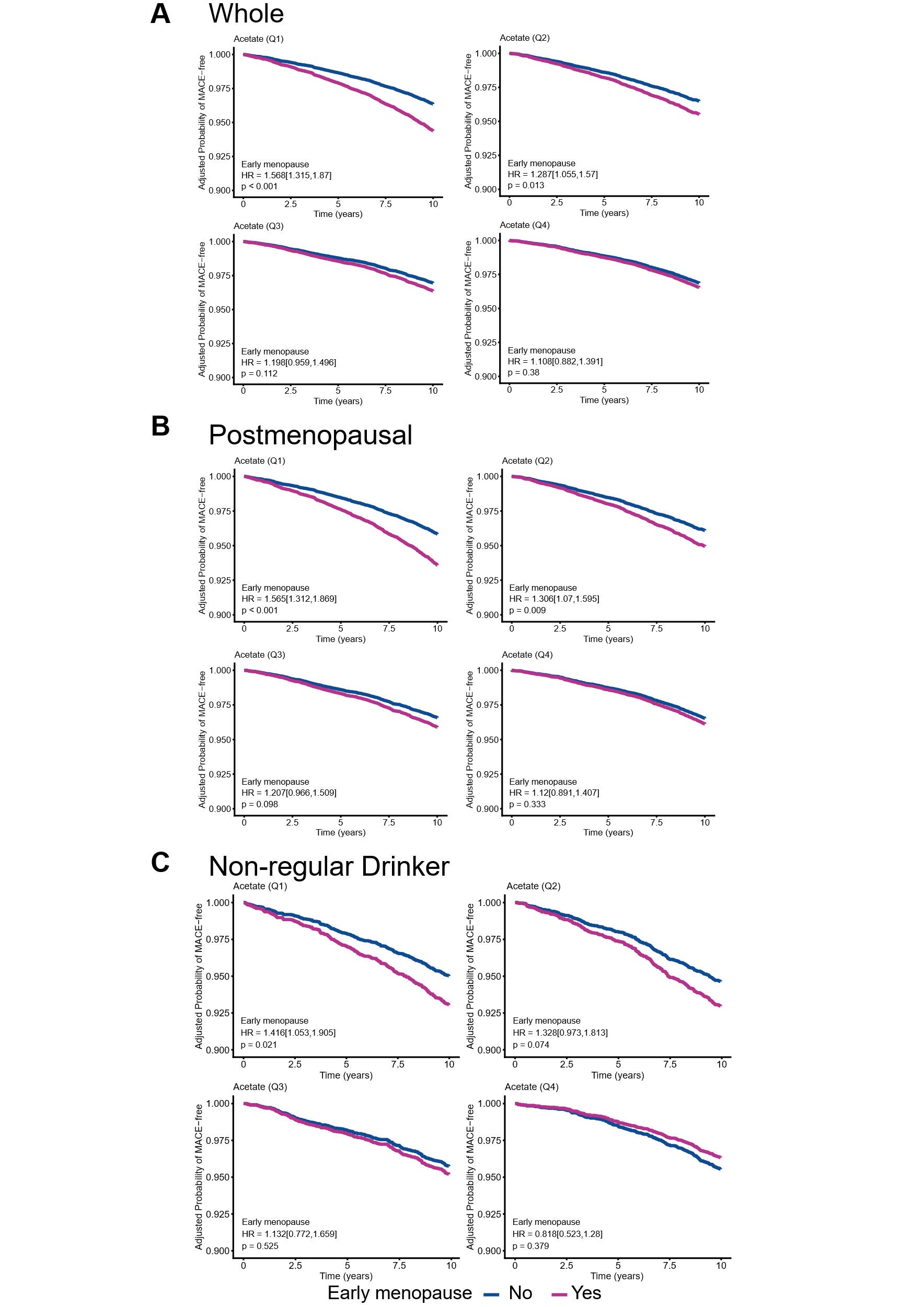

**Figure S3** | **Time-to-event analysis of 10-year MACE incidence associated with early menopause in participants stratified by quantiles of plasma acetate level in UK Biobank** in **A.** the whole cohort (N=81,799), **B.** the postmenopausal cohort (N=68,249), and **C.** non-regular drinkers. (N=19,111). All analyses were adjusted for alcohol consumption, standardised age and BMI, urine potassium and sodium, current and previous HRT use, and also CVD medication intake.

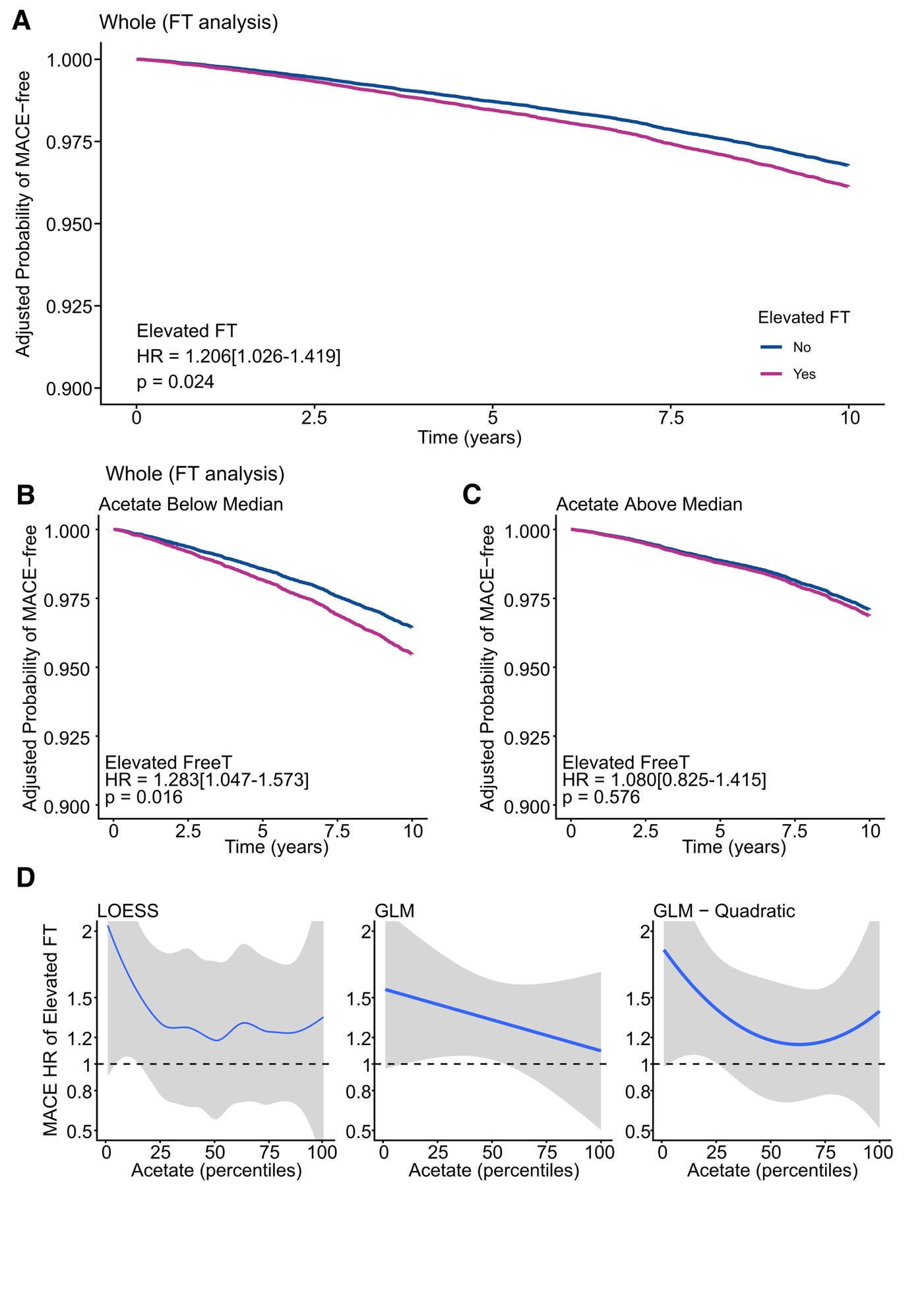

**Figure S4** | **Association of plasma acetate and elevated free testosterone (FT) on 10-year incidence of major adverse cardiovascular events (MACE) in UK Biobank. A.** Time-to-event analysis of adjusted 10-year MACE incidence associated with elevated FT in the analytic cohort of FT (N=59,247). **B-C.** Time-to-event analysis of MACE incidence associated with elevated FT in participants stratified by plasma acetate level (above vs. below median) in the UK Biobank for 10 years in the analytic cohort of FT (N=59,247). **D.** Smoothed curves derived from locally estimated scatterplot smoothing (LOESS), generalised linear regression model (GLM) and GLM including a quadratic term for the adjusted coefficient size of elevated FT across different percentiles of plasma acetate levels (N= 59,247). All analyses were adjusted for alcohol consumption, standardised age and BMI, urine potassium and sodium, TG/HDL-c ratio and also CVD medication intake.
